# Influenza and pneumococcal vaccination and the risk of COVID-19: A systematic review and meta-analysis

**DOI:** 10.1101/2022.06.30.22277089

**Authors:** Georgia G. Kapoula, Konstantina E. Vennou, Pantelis G. Bagos

## Abstract

A number of studies have investigated the potential non-specific effects of some routinely administered vaccines (e.g. influenza, pneumococcal) on COVID-19 related outcomes, with contrasting results. In order to elucidate this discrepancy, we conducted a systematic review and meta-analysis to assess the association between seasonal influenza vaccination and pneumococcal vaccination with SARS-CoV-2 infection and its clinical outcomes. PubMed and medRxiv databases were searched, up until November 2021. Random effects model was used in the meta-analysis to pool odds ratio (OR) and adjusted estimates with their 95% confidence intervals (CIs). Heterogeneity was quantitatively assessed using the Cohran’s *Q* and the *I*^2^ index. Sub-group analysis, sensitivity analysis and assessment of publication bias were performed for all outcomes. In total 38 observational studies were included in the meta-analysis and there was substantial heterogeneity. Influenza and pneumococcal vaccination were associated with lower risk of SARS-Cov-2 infection (OR: 0.80, 95% CI: 0.75-0.86 and OR: 0.70, 95% CI: 0.57-0.88, respectively). Regarding influenza vaccination, it seems that the majority of studies did not properly adjust for all potential confounders, so when the analysis was limited to studies that adjusted for age, sex, comorbidities and socioeconomic indices, the association diminished. This is not the case regarding the pneumococcal vaccination, for which even after adjustment for such factors the association persisted. Regarding harder endpoints such as ICU admission and death, current data do not support the association. Possible explanations are discussed, including trained immunity, inadequate matching for socioeconomic indices and possible coinfection.

## INTRODUCTION

In 2020 in Wuhan, China cases of pneumonia were reported [1], caused by the SARS-CoV-2, which is a coronavirus (CoV) which lead to a pandemic outbreak. The coronaviruses belong in the *Nidovirales* order and in the *Coronaviridae* family, which has four genera divisions, the alpha, beta, gamma, and delta coronaviruses. All coronaviruses contain very large genomes, are enveloped, and non-segmented positive-sense ribonucleic acid (RNA) viruses [2]. Coronaviruses in humans mainly cause respiratory diseases, there are many viruses in this family with a large spectrum of diseases from the less-severe common cold to serious and highly fatal diseases such as the Severe Acute Respiratory Syndrome (SARS) and the Middle East Respiratory Syndrome (MERS). SARS-CoV-2 is a beta coronavirus in the same subgenus with the SARS-CoV-2 and shares RNA with it as well as with the MERS-CoV [3]. The novel coronavirus that we are facing can cause severe disease, a syndrome termed COVID-19. In 1^st^ of April 2022 there have been 486,761,597 confirmed cases and 6,142,735 confirmed deaths worldwide [4].

Seasonal influenza is another acute respiratory infection caused by the influenza viruses. Influenza viruses are part of the *Orthomyxoviridae* family, in which there are three main genera A, B and C [5]. They are RNA viruses with segmented, negative-strand genome. The diseases caused by influenza have a large range from mild to more severe and even death in high□risk groups [6]. As one can see both diseases are respiratory viral diseases, they co-exist at the same period and they can be clinically indistinguishable (fever, cough, nasal congestion or rhinorrhea, myalgia etc.) [7]. Another respiratory infection is pneumonia. The main causes of pneumonia are viruses, bacteria and fungi. One of the most common bacteria which cause pneumonia (mainly in children) is *Streptococcus pneumonia* [8]. Some studies have shown co-infection of COVID-19 and influenza or COVID-19 and pneumonia [9].

One step towards the end of the pandemic is vaccination. There are ten vaccines approved by WHO [10]. Although many non SARS-CoV-2 vaccines have been tested for preventing SARS-CoV-2 infection or having a protective effect in the severity of the disease. The booster Bacillus Calmette-Guérin (BCG) vaccine was suggested, by many studies and even by a meta-analysis, to have beneficial effects on preventing COVID-19 infection [11–13]. Some preliminary studies suggest some protection against SARS-CoV-2 to be conferred from vaccination to other pathogens such as influenza [14] while others have produced contrasting results [15]. In this regard, we conducted a systematic review and meta-analysis to assess the overall association between influenza and pneumococcal vaccination and SARS-CoV-2 infection.

## MATERIALS and METHODS

A comprehensive search was performed to identify papers investigated influenza and pneumococcal vaccination and SARS-CoV-2 infection and severity of the disease. PubMed [16] and medRxiv [17] were investigated for relevant research articles.

The search terms for influenza vaccination were: (”Influenza Vaccines” OR “influenza vaccine” OR “flu vaccine” OR “anti-flu vaccine”) AND (“sars-cov2” OR “covid-19” OR “2019-ncov” OR “2019nconv” OR “sars-cov2” OR “covid-19”) in PubMed and “influenza vaccine” (abstract or title, match all words) in medRxiv. For the pneumococcal vaccination were: (“pneumococcal Vaccines” OR “pneumococcal vaccine”) AND (“sars-cov2” OR “covid-19” OR “2019-ncov” OR “2019nconv” OR “sars-cov2” OR “covid-19”) in PubMed and “pneumococcal vaccine” (abstract or title, match all words) in medRxiv. To avoid selection bias no filter was applied in language, year, method etc. This systematic review was performed according to Preferred Reporting Items for Systematic Reviews and Meta-Analyses (PRISMA) guidelines [18], the STROBE Statement checklist [19] and other general guidelines [20]. The search was performed until 01/11/2021.

The information that was extracted from the selected studies was: Authors, Title, Year of publication, Country in which it was conducted, Study Design, Study population, Sample size, Gender, Comorbidities, COVID-19 outcome, the effect size related to relevant outcomes (e.g. OR, HR, RR) and other potentially relevant information. Since the risk for COVID-19 varies largely according to risk factors, studies not including adjusted odds ratio (OR), adjusted hazard ratio (HR) or adjusted relative risk (RR) as their effect size were excluded. We requested that the effect sizes were adjusted mainly for gender, age, race, comorbidities, smoking status etc.

The main inclusion criteria were: a) vaccination period for influenza or pneumococcal from autumn 2019 until autumn 2020, b) confirmed COVID-19 infection with either real-time PCR, or serologic test, or according to WHO definition or laboratory confirmation (not specified), c) participants being adults and d) comparison of people infected with COVID-19 against non-infected ones using an adjusted estimate of OR, HR or RR (see above).

In order to assess the association between the two vaccines (influenza and pneumococcal) and COVID-19 outcome (infection, death, hospitalization, etc.), a random effects meta-analysis was performed, for each outcome, using as effect size the adjusted OR, or adjusted RR/HR, combined with their 95% confidence interval (CI) [21]. The between studies heterogeneity of the pooled estimates was quantified by means of the chi-square-based Cochran’s *Q* statistic and the consistency index (*I^2^*) [22]. To assess publication bias, the rank correlation method of Begg and Mazumdar [23], and the fixed effects regression method of Egger [24] were applied. Subgroup analyses were performed to investigate the effect of various study level characteristics. Sensitivity analyses were performed by omitting one study at a time to assess studies with notable impact and examine the robustness of the overall effect. For all statistical analyses performed, the statistical software package STATA 13 [25] was used and results with *p*-value <0.05 were considered statistically significant.

## RESULTS

### Study selection and study characteristics

The literature search from the two databases identified a total of 768 related to the influenza vaccination and 86 records for the pneumococcal vaccination search. After reading the titles for excluding duplicates, 674 and 79 unique publications remained for influenza and pneumococcal vaccination, respectively. The simultaneously titles and abstracts review led to a further rejection of 697 publications. The remaining 56 articles were read in full copy, as well as the citations of the retrieved articles. Finally, 38 publications met all the inclusion criteria for the quantitative analysis and were included in the final systematic review and meta-analysis. The PRISMA flow diagram of the study selection procedure is shown in Figure 1.

**Figure 1.**
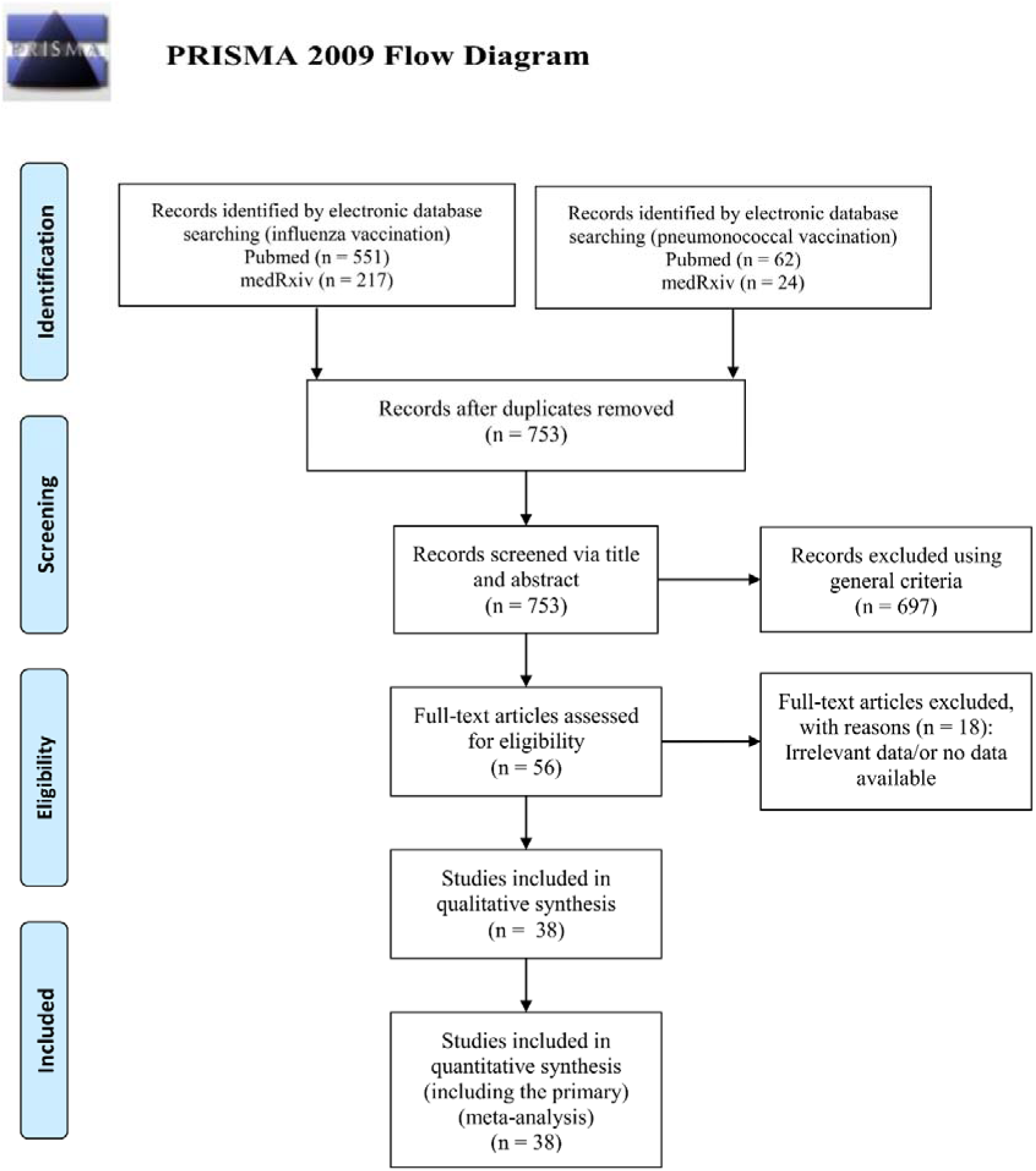
The PRISMA flow diagram for the included studies

Among the included publications, 22 studies focused on the association between influenza vaccination and the risk of SARS-CoV-2 infection and 10 studies on the association between pneumococcal vaccination and the risk of SARS-CoV-2 infection. These studies comprised a total of 55,917,587 individuals (COVID-19 patients and non-COVID patients).

Out of the 38 publications included in the meta-analysis, 19 publications assessed the association of the influenza vaccination and COVID-19 clinical outcomes. The different outcomes included: a) need of hospitalization involving 15 studies with 55,881,321 individuals (13,123,605 vaccinated and 42,758,116 unvaccinated patients) b) administration of mechanical ventilation/invasive respiratory oxygen support described in 4 studies encompassing 255,735 patients, c) intensive care unit admission. This outcome was assessed in 11 studies involving 446,924 vaccinated and not vaccinated patients and d) mortality which was assessed in 18 studies involving 407,409 vaccinated and not vaccinated COVID-19 patients.

Moreover, 4 studies assessed the association between pneumococcal vaccination and hospitalization enrolling 4,310 patients of which 1,333 were vaccinated and 2,977 were not vaccinated. Finally, 3 studies examined the association between pneumococcal vaccination and intensive care unit involving 1,231 patients (921 non-vaccinated Vs 310 vaccinated). The baseline characteristics of the included studies are listed in Table 1 (influenza vaccination and SARS-CoV-2 infection), Table 2 (influenza vaccination and clinical outcomes) and Τable 3 (pneumococcal vaccination and SARS-CoV-2 infection and clinical outcomes). Most studies enrolled individuals from the general population. Among them, five studies had as a population study health workers [26–30], one study involved only pregnant women [31], another study included only transplant recipients [32] and four studies were performed in older adults (age >65) [33–36]. The laboratory method (rt-PCR) used to diagnose of the disease was common in most of the included studies. The adjustment variables varied across studies with age, sex and comorbidities being the most common ones reflecting the known COVID-19 risk factors. Studies which did not contain adjusted estimates (OR or RR/HR) as well as ecological studies were excluded from the meta-analysis and are summarized in Supplementary Τable 1.

**Table 1.** Baseline characteristics of the 21 publications, involving 22 studies that assessed the association between influenza vaccination and SARS-CoV-2 infection.

| First Author's name | Year of publication | Country | study design/study population | Identification of COVID-19 | Sample size | Effect size | Adjusted estimate | Adjusted factors |
| --- | --- | --- | --- | --- | --- | --- | --- | --- |
| M. Noale et al. [35] | 2020 | Italy | Cross-sectional study/Adults-general population | rt-PCR | 6,061 | OR | 0.85 (95% CI: 0.74-0.98) | age, sex, self-reported comorbidities, education, area of residence, smoking status |
| M. Noale et al. [35] | 2020 | Italy | Cross-sectional study/Old adults | rt-PCR | 619 | OR | 0.87 (95% CI: 0.59-1.28) | sex, age, self-reported comorbidities, education, area of residence, smoking status |
| I. Martínez-Baz et al. [26] | 2020 | Spain | Prospective cohort study/Health workers | rt-PCR or Antibody Rapid test | 9,745 | OR | 1.07(95% CI: 0.92-1.24) | age, sex, major chronic conditions, profession, and any ILI diagnosis in the previous five years. |
| P. Ragni et al. [60] | 2020 | Italy | Case control study/Adults-general population | rt-PCR | 17,608 | OR | 0.89 (95% CI: 0.80-0.99) | age, sex, Charlson index score and time of the swab test |
| M. Belingheri et al. [28] | 2020 | Italy | Retrospective cohort study/Health workers | rt-PCR | 3,520 | OR | 0.41 (95% CI: 0.07-2.39) | age, gender, number of flu vaccination uptakes |
| A. Vila-Córcoles et al. [61] | 2020 | Spain | Retrospective cohort study/Adults-general population | rt-PCR | 79,083 | HR | 1.02 (95% CI: 0.79-1.32) | age, sex, comorbidities, residence and vaccination history |
| I. Green et al. [62] | 2020 | Israel | Retrospective cohort study/Adults-general population | rt-PCR | 19,089 | OR | 0.79 (95% CI: 0.67-0.98) | age, gender, ethnicity, underlying medical conditions, obesity, and smoking status |
| E. Kissling et al. [15] | 2021 | Europe | Case-control study (multicenter)/Adults-general population | rt-PCR | 2,147 | OR | 0.93 (95% CI: 0.66-1.32) | age, sex and chronic conditions, time of swab test |
| A. Conlon et al. [63] | 2021 | USA | Retrospective cohort study/Adults-general population | rt-PCR | 27,201 | OR | 0.76 (95% CI: 0.68-0.86) | age, sex, ethnicity, race, BMI, Elixhauser score, comorbidities, smoking status |
| B. Erismis et al. [30] | 2021 | Turkey | Retrospective cohort study/Health workers | Not specified | 203 | RR | 0.83 (95% CI: 0.75-0.93) | sex |
| A. Bozek et al. [64] | 2021 | Poland | Retrospective cohort study (multicenter)/Adults-general population | rt-PCR | 2,558 | HR | 0.74 (95% CI: 0.54-0.89) | age, sex, comorbidities, high economic status, BCG vaccination in the past |
| M. Kowalska et al. [65] | 2021 | Poland | Cross-sectional study/Adults-general population | IgG antibodies | 5,376 | OR | 0.68 (95% CI: 0.55-0.83) | age, sex, comorbidities, previous COVID-19 contact or quarantine |
| M. Fernández-Prada et al. [66] | 2021 | Spain | Case-control study/Adults-general population | rt-PCR | 188 | OR | 1.70 (95% CI: 0.97-3.25) | age, sex, patient location |
| K. Huang et al. [34] | 2021 | USA | Cross-sectional study/Old adults | Not specified | 55,667,997 | OR | 0.76 (95% CI: 0.75-0.77) | age, sex, comorbidities |
| J. P. King et al. [67] | 2021 | USA | Prospective cohort study/Adults-general population | rt-PCR | 1,356 | OR | 0.83 (95% CI: 0.63-1.10) | age, sample collection interval, sex, high-risk condition and month of onset |
| C. Pawlowski et al. [68] | 2021 | USA | Retrospective cohort study/Adults-general population | rt-PCR | 12,791 | RR | 0.85 (95% CI: 0.75-0.96) | age, gender, race, ethnicity, county level COVID-19 incidence rate, county-level COVID-19 test positive rate, Elixhauser comorbidities, pregnancy, number of other vaccines |
| S. Caratozzolo et al. [33] | 2020 | Italy | Retrospective cohort study/Old adults | rt-PCR/or infection according to WHO definition | 848 | OR | 0.47 (95% CI: 0.29-0.74) | age, sex, comorbidities, clinical dementia rating scale |
| M. N. Rivas et al. [29] | 2021 | USA | Retrospective cohort study/Health workers | IgG antibodies | 6,087 | OR | 1.84 (95% CI: 0.57-11.3) | age, sex |
| J. G. Zein et al. [69] | 2020 | USA | Prospective cohort study/Adults-general population | Not specified | 13,220 | OR | 0.79 (95% CI: 0.62-1.00) | age, sex, race and comorbidities |
| P. A. Debisarun et al. [37] | 2020 | Netherlands | Retrospective cohort study/Health workers | rt-PCR | 6,856 | RR | 0.61 (95% CI: 0.46-0.82) | age |
| Y. Xiang et al. [70] | 2020 | UK | Prospective cohort study/Adults-general population | Laboratory confirmed | 30,835 | OR | 0.60 (95% CI: 0.53-0.68) | age, sex, ethnic group, comorbidities, socioeconomic status, smoking status, anthropometric measures |
| M. Alkathlan et al. [71] | 2021 | Saudi Arabia | Cross-sectional study/Health workers | Laboratory confirmed | 424 | OR | 0.83 (95% CI: 0.51-1.35) | age, sex, comorbidities, nationality, educational level, flu vaccine efficacy |

OR=adjusted odds ratio, HR=hazard odds ratio, RR=relative risk, rt-PCR=Reverse transcription polymerase chain reaction

**Table 2.** Baseline characteristics of the 17 publications involving studies that assessed the association between influenza vaccination and COVID-19clinical outcomes: (15 studies for hospitalization, 4 for Mechanical Ventilation/invasive respiratory support, 11 for Intensive care and 18 for mortality).

| First Author's name | Year of publication | Country | Study design | Identification of COVID-19 | Sample size | Effect size | Adjusted estimate | Adjusted factors |
| --- | --- | --- | --- | --- | --- | --- | --- | --- |
| **HOSPITALIZATION** |  |  |  |  |  |  |  |  |
| P. D. Pedote et al. [72] | 2021 | Italy | Retrospective cohort study | rt-PCR | 662 | OR | 1.20 (95% CI: 0.70-1.90) | age, sex, chronic disease |
| R. Pastorino et al. [36] | 2021 | Italy | Prospective cohort study | rt-PCR | 741 | OR | 1.03 (95% CI: 0.66-1.62) | age, sex, comorbidities |
| C. Pawlowski et al. [68] | 2021 | USA | Retrospective cohort study | rt-PCR | 959 | RR | 1.10 (95% CI: 0.83-1.50) | age, gender, race, ethnicity, county level COVID-19 incidence rate, county-level COVID-19 test positive rate, Elixhauser comorbidities, pregnancy, number of other vaccines |
| M. Gobbato et al. [73] | 2020 | Italy | Retrospective cohort study | Laboratory confirmed | 3,010 | OR | 0.78 (95% CI: 0.61-1.01) | age, sex, comorbidities, medication, health district |
| M-J. Yang et al. [74] | 2020 | USA | Retrospective cohort study (single-centered) | Laboratory confirmed | 2,005 | OR | 0.41 (95% CI: 0.28-0.59) | age, sex, ethnicity/race, comorbidities |
| S. Greco et al. [38] | 2021 | Italy | Retrospective cohort study (multicenter) | rt-PCR | 952 | OR | 1.44 (95% CI: 1.01-2.05) | sex |
| A. Bozek et al. [64] | 2021 | Poland | Retrospective cohort study (multicenter) | rt-PCR | 151 | HR | 0.48 (95% CI: 0.36-0.77) | age, sex, comorbidities, high economic status, BCG vaccination in the past |
| K. Huang et al. [34] | 2021 | USA | Cross-sectional study | Not specified | 55,667,977 | OR | 0.76 (95% CI: 0.75-0.77) | age, sex, comorbidities |
| M. Massari et al. [75] | 2021 | Italy | Retrospective cohort study | rt-PCR | 115,945 | RR | 0.87 (95% CI: 0.86-0.88) | age, sex, Charlson comorbidity index, ethnicity, drugs, comorbidities |
| A. Conlon et al. [63] | 2021 | USA | Retrospective cohort study | rt-PCR | 1,218 | OR | 0.58 (95% CI: 0.46-0.73) | age, sex, ethnicity, race, BMI, Elixhauser score, comorbidities, smoking status |
| P. Ragni et al. [60] | 2020 | Italy | Prospective cohort study | rt-PCR | 4,485 | HR | 1.00 (95% CI: 0.84-1.19) | age, comorbidities, exposure to influenza vaccination |
| C. R. Wilcox et al. [76] | 2021 | UK | Retrospective cohort study | rt-PCR | 6,921 | HR | 0.85(95% CI: 0.75-0.97) | age, sex, BMI, socioeconomic status (IMD), smoking status, frailty score (electronic frailty index), pre-existing comorbidities and medication |
| J. G. Zein et al. [69] | 2020 | USA | Prospective cohort study | Not specified | 1,434 | OR | 1.29 (95% CI: 0.72-2.31) | age, sex, race and comorbidities |
| I. Ilic et al. [77] | 2020 | Serbia | Retrospective cohort study | rt-PCR | 107 | OR | 1.31 (95% CI: 0.54-3.17) | age, malignancy, hypertension, coronary artery disease, heart failure, diabetes mellitus, bronchial asthma, previous CVD, arrhythmia, BMI, current smoking |
| S. M. Taghioff et al. [78] | 2021 | Netherlands | Retrospective cohort study | Not specified | 74,754 | OR | 1.07 (95% CI: 0.96-1.18) | age, race, ethnicity, gender, hypertension, diabetes, hyperlipidemia, chronic obstructive pulmonary disease (COPD), obesity, heart disease, and lifestyle habits such as smoking |
| **MECHANICAL VENTILATION/INVANSIVE RESPIRATORY SUPPORT** |  |  |  |  |  |  |  |  |
| G. Fink et al. [79] | 2020 | Brazil | Retrospective cohort study | rt-PCR | 39,745 | OR | 0.83 (95% CI: 0.77-0.89) | age, sex, race, educational attainment, comorbidities, treatment facility |
| A. Conlon et al. [63] | 2021 | USA | Retrospective cohort study | rt-PCR | 1,218 | OR | 0.45 (95% CI: 0.27-0.78) | age, sex, ethnicity, race, BMI, Elixhauser score, comorbidities, smoking status |
| A. E. Demkina et al. [80] | 2020 | Russia | Cohort study | Clinically confirmed/<br>rt-PCR | 214,751 | OR | 0.74 (95% CI: 0.54-1.01) | age, patient's gender, comorbidities |
| A. Bozek et al. [64] | 2021 | Poland | Retrospective cohort study (multicenter) | rt-PCR | 21 | OR | 0.42 (95% CI: 0.02-9.96) | age , sex, comorbidities, high economic status, BCG vaccination in the past |
| **INTENSIVE CARE** |  |  |  |  |  |  |  |  |
| M-J. Yang et al. [74] | 2021 | USA | Retrospective cohort study (single-centered) | rt-PCR | 2,005 | OR | 0.31 (95% CI: 0.07-0.85) | age, sex, ethnicity/race, comorbidities |
| M. Massari et al. [75] | 2021 | Italy | Retrospective cohort study | rt-PCR | 111,740 | RR | 1.01 (95% CI: 0.99-1.04) | age, sex, comorbidities, ethnicity, drugs |
| R. Pastorino et al. [36] | 2021 | Italy | Prospective cohort study | rt-PCR | 99 | OR | 1.26 (95% CI: 0.74-2.21) | age, sex, comorbidities |
| A. Conlon et al. [63] | 2021 | USA | Retrospective cohort study | rt-PCR | 1,218 | OR | 0.64 (95% CI: 0.41-1.00) | age, sex, ethnicity, race, BMI, Elixhauser score, comorbidities, smoking status |
| C. Pawlowski et al. [68] | 2021 | USA | Retrospective cohort study | rt-PCR | 959 | RR | 1.10 (95% CI: 0.56-2.20) | age, gender, race, ethnicity, county level COVID-19 incidence rate, county-level COVID-19 test positive rate, Elixhauser comorbidities, pregnancy, number of other vaccines |
| G. Fink et al. [79] | 2020 | Brazil | Retrospective cohort study | rt-PCR | 39,156 | OR | 0.93 (95% CI: 0.87-0.99) | age, sex, race, educational attainment, comorbidities, treatment facility |
| A. E. Demkina et al. [80] | 2020 | Russia | Retrospective cohort study | Clinically confirmed/<br>rt-PCR | 214,751 | OR | 0.76 (95% CI: 0.59-0.97) | age, patient's gender, comorbidities |
| M. Candelli et al. [81] | 2021 | Italy | Retrospective cohort study | rt-PCR | 602 | OR | 0.73 (95% CI: 0.35-1.56) | age, sex, comorbidities |
| M. L. de la Cruz Conty et al. [31] | 2021 | Spain | Prospective cohort study (multicenter) | rt-PCR | 206 | OR | 1.92 (95% CI: 0.36-10.3) | age, sex, comorbidities |
| J. G. Zeinet al. [69] | 2020 | USA | Prospective cohort study | Not specified | 1,434 | OR | 0.65 (95% CI: 0.22-1.79) | age, sex, race and comorbidities |
| S. M. Taghioff et al. [78] | 2021 | Netherlands | Retrospective cohort study | Not specified | 74,754 | OR | 1.18 (95% CI: 1.00-1.39) | age, race, ethnicity, gender, hypertension, diabetes, hyperlipidemia, chronic obstructive pulmonary disease (COPD), obesity, heart disease, and lifestyle habits such as smoking |
| **MORTALITY** |  |  |  |  |  |  |  |  |
| S. Greco et al. [38] | 2021 | Italy | Retrospective cohort study | rt-PCR | 952 | OR | 1.06 (95% CI: 0.60-1.88) | sex |
| P. D. Pedote et al. [72] | 2021 | Italy | Retrospective cohort study | rt-PCR | 662 | OR | 1.60 (95% CI: 0.80-3.20) | age, sex, chronic diseases |
| R. Pastorino et al. [36] | 2021 | Italy | Prospective cohort study | rt-PCR | 97 | OR | 1.33 (95% CI: 0.77-2.31) | age, sex, comorbidities |
| M. Massari et al. [75] | 2021 | Italy | Retrospective cohort study | rt-PCR | 115,945 | RR | 1.04 (95% CI: 1.01-1.06) | age, sex, comorbidities, ethnicity, drugs |
| J. M. Fernadez. Ibáñez et al. [82] | 2021 | Spain | Retrospective cohort study | rt-PCR | 410 | OR | 1.55 (95% CI: 0.96-2.48) | sex, chronic diseases |
| A. Bozek et al. [64] | 2021 | Poland | Retrospective cohort study (multicenter) | rt-PCR | 2,558 | HR | 0.74 (95% CI: 0.03-20.8) | age, sex, comorbidities, high economic status, BCG vaccination in the past |
| M. Candelli et al. [81] | 2021 | Italy | Retrospective cohort study | rt-PCR | 602 | OR | 0.20 (95% CI: 0.08-0.51) | age, sex, comorbidities |
| A. Conlon et al. [63] | 2021 | USA | Retrospective cohort study | rt-PCR | 1,218 | HR | 0.94 (95% CI: 0.61-1.47) | age, sex, ethnicity, race, BMI, Elixhauser score, comorbidities, smoking status |
| P. Ragni et al. [60] | 2020 | Italy | Prospective cohort study | rt-PCR | 4,872 | HR | 1.14 (95% CI: 0.95-1.37) | age, comorbidities, exposure to influenza vaccination test |
| C. R Wilcox et al. [76] | 2021 | UK | Retrospective cohort study | rt-PCR | 6,368 | OR | 0.76 (95% CI: 0.64-0.90) | age, sex, BMI, socioeconomic status (IMD), smoking status, frailty score (electronic frailty index), pre-existing comorbidities and medication |
| G. Fink et al. [79] | 2020 | Brazil | Retrospective cohort study | rt-PCR | 53,752 | OR | 0.84 (95% CI: 0.78-0.91) | age, sex, race, educational attainment, comorbidities, treatment facility |
| A. E. Demkina et al. [80] | 2020 | Russia | Retrospective cohort study | Clinically confirmed/<br>rt-PCR | 117,346 | HR | 0.78 (95% CI: 0.63-0.95) | age, patient's gender, comorbidities |
| Y. Azzi et al. [32] | 2020 | USA | Prospective cohort study | rt-PCR | 229 | OR | 0.88 (95% CI: 0.70-0.96) | age, type of kidney transplant |
| M. Gobbato et al. [73] | 2020 | Italy | Retrospective cohort study | Laboratory confirmed | 3,010 | OR | 0.78 (95% CI: 0.61-1.01) | age, sex, comorbidities, medication, health district |
| J. G. Zeinet al. [69] | 2020 | USA | Prospective cohort study | Not specified | 14,654 | OR | 0.98 (95% CI: 0.39-2.43) | age, sex, race and comorbidities |
| D. Giannoglou et al. [83] | 2020 | Greece | Cross-sectional study | Not specified | 512 | OR | 0.38 (95% CI: 0.17-0.81) | age, gender, comorbidities |
| E. Ortiz-Prado et al. [84] | 2020 | Ecuador | Cross-sectional study | rt-PCR | 9,468 | RR | 1.40 (95% CI: 0.46-4.28) | age, gender, comorbidities |
| S. M. Taghioff et al [78] | 2021 | Netherlands | Retrospective cohort study | Not specified | 74,754 | OR | 0.89 (95% CI: 0.77-1.03) | age, race, ethnicity, gender, hypertension, diabetes, hyperlipidemia, chronic obstructive pulmonary disease (COPD), obesity, heart disease, and lifestyle habits such as smoking. |

### The association between influenza vaccination and SARS-CoV-2 infection

The results of the random effects analysis are summarized in Table 4. The association of the influenza vaccination and SARS-CoV-2 infection is presented graphically in the forest plot in Figure 2. Influenza vaccination is shown to be associated with lower risk of SARS-CoV-2 infection (random effects model: pooled estimate 0.80, 95% CI: 0.75-0.86). The results of the statistical tests for publication bias (Begg’s, Egger’s test) are shown in Table 4. Both tests showed no evidence of publication bias for the pooled estimates of the association between influenza vaccination and SARS-CoV-19 infection (*p*-value >0.05). In the leave-one-out sensitivity analyses regarding the association between influenza vaccination and SARS-CoV-2 infection, the results showed that no individual study influenced the overall effect estimate.

**Figure 2.**
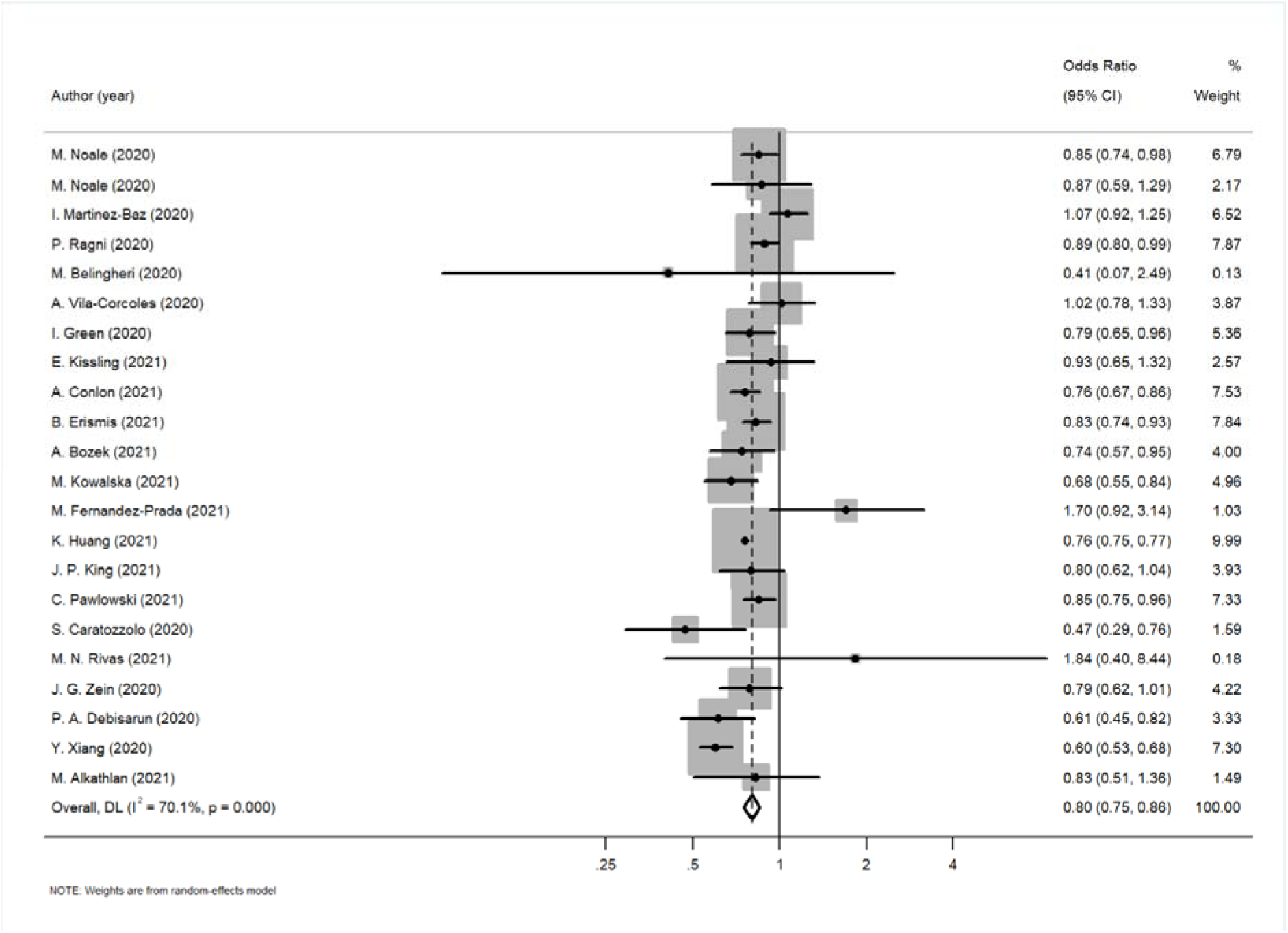
Forest plot for the association between influenza vaccination and SARS-Cov-2 infection

**Table 3.** Baseline characteristics of a) 6 publications, involving 10 studies, that assessed the association between pneumococcal vaccination and SARS-CoV-2 infection, b) 4 studies that assessed that assessed the association between pneumococcal vaccination and the risk of hospitalization and c) 3 studies that assessed that assessed the association between pneumococcal vaccination and the risk of intensive care.

| First Author's name | Year of publication | Country | Study design | Identification of COVID-19 | Sample size | Effect size | Adjusted estimate | Adjusted factors |
| --- | --- | --- | --- | --- | --- | --- | --- | --- |
| **INFECTION** |  |  |  |  |  |  |  |  |
| A. Vila-Córcoles et al. [61] | 2020 | Spain | Retrospective cohort study | rt-PCR | 79,083 | HR | 1.02 (95% CI 0.78-1.33) | age, sex, comorbidities, residence and vaccination history |
| M. Noale et al. [35] | 2020 | Italy | Cross-sectional study | rt-PCR | 6,061 | OR | 0.61 (95% CI 0.41-0.91) | sex, age, comorbidities, education, area of residence, smoking status |
| M. Noale et al. [35] | 2020 | Italy | Cross-sectional study (>65 years) | rt-PCR | 619 | OR | 0.56 (95% CI 0.33-0.95) | sex, age, comorbidities, education, area of residence, smoking status |
| C. Pawlowski et al. [68] | 2021 | USA | Retrospective cohort study | rt-PCR | 4,693 | RR | 0.72 (95% CI 0.56-0.92) | age, gender, race, ethnicity, county level COVID-19 incidence rate, county-level COVID-19 test positive rate, Elixhauser comorbidities, pregnancy, number of other vaccines |
| C. Pawlowski et al. [68] | 2021 | USA | Retrospective cohort study | rt-PCR | 4,636 | RR | 1.06 (95% CI 0.81-137) | age, gender, race, ethnicity, county level COVID-19 incidence rate, county-level COVID-19 test positive rate, Elixhauser comorbidities, pregnancy, number of other vaccines |
| M. N. Rivas et al. [29] | 2021 | USA | Retrospective cohort study | IgG antibodies | 6,083 | OR | 0.99 (95% CI 0.71-1.36) | age, sex |
| M. Fernández-Prada et al. [66] | 2021 | Spain | Case-control study | rt-PCR | 188 | OR | 0.40 (95% CI 0.17-1.01) | age, sex and patient location |
| M. Fernández-Prada et al. [66] | 2021 | Spain | Test-negative case-control study | rt-PCR | 187 | OR | 0.70 (95% CI: 0.28-2.10) | age, sex and patient location |
| M. Fernández-Prada et al. [66] | 2021 | Spain | Case-control study | rt-PCR | 188 | OR | 0.20 (95% CI: 0.06-1.18) | age, sex and patient location |
| Y. Xiang et al. [70] | 2020 | UK | Prospective cohort study | Laboratory confirmed | 30,835 | OR | 0.5 (95% CI: 0.31-0.82) | age, sex, ethnic group, comorbidities, socioeconomic status, smoking status, anthropometric measures |
| **HOSPITALIZATION** |  |  |  |  |  |  |  |  |
| M. Gobbato et al. [73] | 2020 | Italy | Retrospective cohort | Laboratory confirmed | 3,010 | OR | 1.53 (95% CI: 1.91-1.97) | age, sex, comorbidities, medication, health district |
| R. Pastorino et al. [36] | 2021 | Italy | Prospective cohort study | rt-PCR | 741 | OR | 0.96 (95% CI: 0.53-1.78) | age, sex, comorbidities |
| C. Pawlowski et al. [68] | 2021 | USA | Retrospective cohort study | rt-PCR | 277 | RR | 1.30 (95% CI: 0.86-2.10) | age, gender, race, ethnicity, county level COVID-19 incidence rate, county-level COVID-19 test positive rate, Elixhauser comorbidities, pregnancy, number of other vaccines |
| C. Pawlowski et al. [68] | 2021 | USA | Retrospective cohort study | rt-PCR | 282 | RR | 1.20 (95% CI: 0.71-2.20) | age, gender, race, ethnicity, county level COVID-19 incidence rate, county-level COVID-19 test positive rate, Elixhauser comorbidities, pregnancy, number of other vaccines |
| **INTENSIVE CARE UNIT** |  |  |  |  |  |  |  |  |
| R. Pastorino et al. [36] | 2021 | Italy | Prospective cohort study | rt-PCR | 741 | OR | 0.75 (95% CI: 0.35-1.61) | age, sex, comorbidities |
| C. Pawlowski et al. [68] | 2021 | USA | Retrospective cohort study | rt-PCR | 254 | RR | 1.40 (95% CI: 0.48-3.90) | vaccinated and unvaccinated propensity score matching; demographics (age, gender, race, ethnicity), county level COVID-19 incidence rate, county-level |
|  |  |  |  |  |  |  |  | COVID-19 test positive rate, Elixhauser comorbidities, pregnancy, number of other vaccines |
| C.Pawlowski et al. [68] | 2021 | USA | Retrospective cohort study | rt-PCR | 236 | RR | 1.10 (95% CI: 0.46-2.40) | age, gender, race, ethnicity, county level COVID-19 incidence rate, county-level COVID-19 test positive rate, Elixhauser comorbidities, pregnancy, number of other vaccines |

**Table 4.**
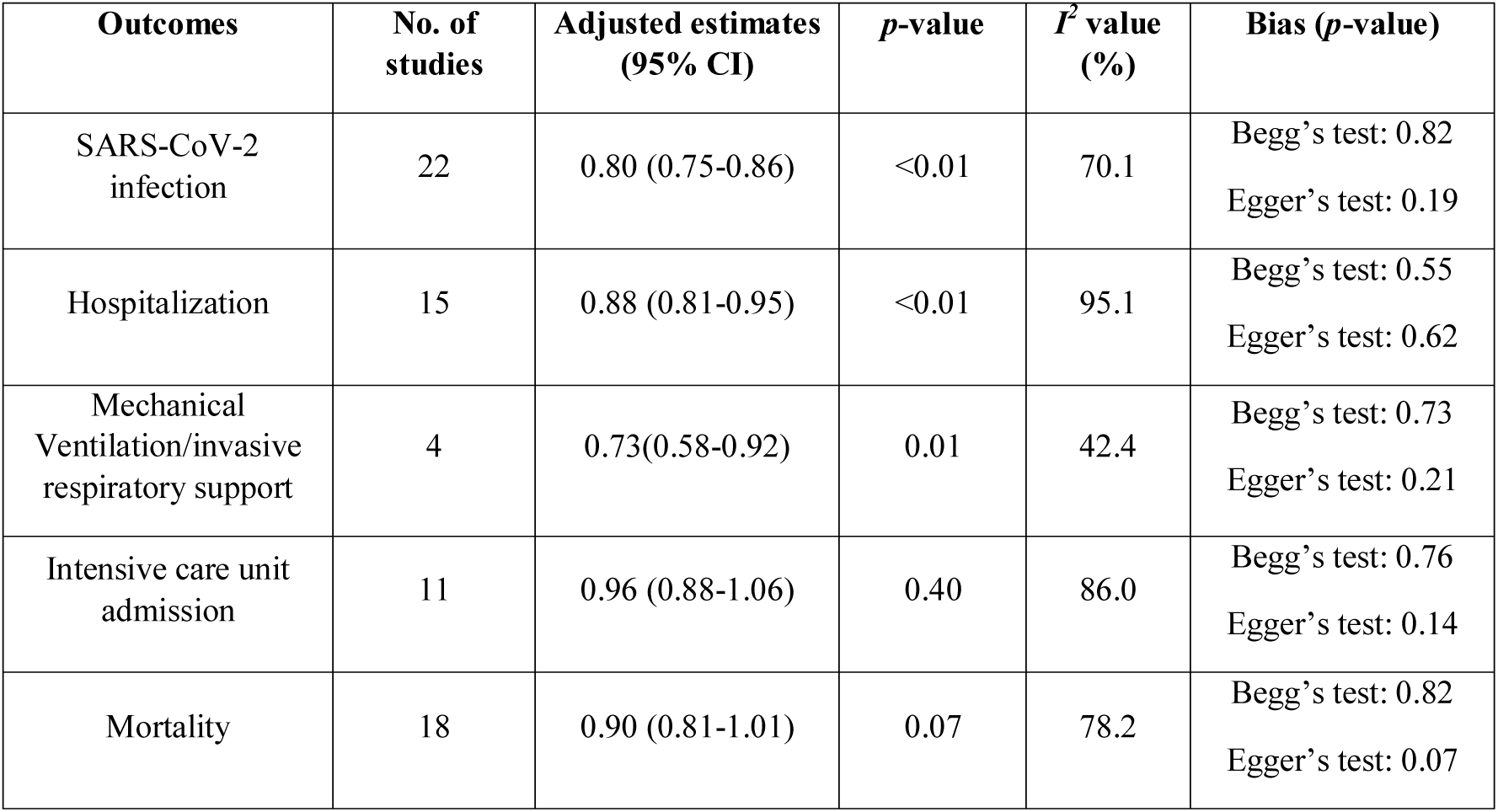
Effect sizes, heterogeneity statistics and results for publications bias of the association between influenza vaccination, SARS-CoV-2 infection as well as clinical outcomes.

| **Outcomes** | **No. of studies** | **Adjusted estimates (95% CI)** | ***p*-value** | ***I*<sup>2</sup> value (%)** | **Bias (*p*-value)** |
| --- | --- | --- | --- | --- | --- |
| SARS-CoV-2 infection | 22 | 0.80 (0.75-0.86) | <0.01 | 70.1 | Begg's test: 0.82<br>Egger's test: 0.19 |
| Hospitalization | 15 | 0.88 (0.81-0.95) | <0.01 | 95.1 | Begg's test: 0.55<br>Egger's test: 0.62 |
| Mechanical Ventilation/invasive respiratory support | 4 | 0.73(0.58-0.92) | 0.01 | 42.4 | Begg's test: 0.73<br>Egger's test: 0.21 |
| Intensive care unit admission | 11 | 0.96 (0.88-1.06) | 0.40 | 86.0 | Begg's test: 0.76<br>Egger's test: 0.14 |
| Mortality | 18 | 0.90 (0.81-1.01) | 0.07 | 78.2 | Begg's test: 0.82<br>Egger's test: 0.07 |

The between-studies heterogeneity was substantial with *I^2^*=70.1%. In order to investigate the high degree of heterogeneity we performed subgroup analysis investigating the potential effect of study-level characteristics (study design, study population, effect size, adjustment factors, method for assessment of COVID-19 and continent). However, none of these factors seemed to play an important role, since in all cases the results of the subgroup analysis are quite close to the overall estimate (Table 5). It is worth noting however, that the association was diminished when the analysis was restricted to the 6 studies that adjusted for age, sex, comorbidities and some indices of socioeconomic status (like residence, income or education).

**Table 5.** Subgroup analysis of the association between influenza vaccination and SARS-CoV-2 infection.

| Subgroup variables | No. of studies | Adjusted estimate (95% CI) | p-value | I<sup>2</sup> value (%) |
| --- | --- | --- | --- | --- |
| **Study Design** |  |  |  |  |
| cross sectional study | 5 | 0.76 (0.75-0.77) | <0.01 | 0.00 |
| retrospective cohort study | 10 | 0.80 (0.75-0.86) | <0.01 | 41.5 |
| prospective cohort study | 4 | 0.80 (0.56-1.08) | 0.14 | 90.9 |
| case-control study | 3 | 0.99 (0.76-1.23) | 0.93 | 51.7 |
| **Study Population** |  |  |  |  |
| adults-general population | 13 | 0.80 (0.74-0.87) | <0.01 | 68.4 |
| older adults | 3 | 0.72 (0.56-0.92) | 0.01 | 53.8 |
| Health workers | 6 | 0.84 (0.68-1.04) | 0.12 | 66.6 |
| **Effect Size** |  |  |  |  |
| OR | 17 | 0.80 (0.74-0.87) | <0.01 | 72.5 |
| RR/HR | 5 | 0.82 (0.73-0.92) | <0.01 | 46.4 |
| **Adjustment Factors** |  |  |  |  |
| age/sex/age and sex | 5 | 0.86 (0.62-1.21) | 0.40 | 63.5 |
| age, sex and comorbidities | 9 | 0.82 (0.73-0.92) | <0.01 | 78.4 |
| age, sex, comorbidities, and socioeconomic status, education or residence | 6 | 0.87 (0.68-1.11) | 0.26 | 81.8 |
| **COVID-19 test** |  |  |  |  |
| rt-PCR | 15 | 0.84 (0.77-0.92) | <0.01 | 57.7 |
| IgG/IgM antibodies | 2 | 0.83 (0.38-1.82) | 0.64 | 37.9 |
| Laboratory confirmed (not specified) | 5 | 0.74 (0.66-0.83) | <0.01 | 74.9 |
| **Continent** |  |  |  |  |
| Europe | 13 | 0.81 (0.70-0.93) | <0.01 | 79.7 |
| N. America | 6 | 0.76 (0.75-0.77) | <0.01 | 0.00 |
| Asia | 3 | 0.83 (0.77-0.90) | <0.01 | 0.00 |

Moreover, two studies (Debisarum et al. [37], Erismis et al [30]) were scrutinized further as in their analysis they used only age and sex, respectively as adjustment variables. However, when both were excluded from the meta-analysis and the overall meta-analysis was repeated, the results did not differ from the overall estimate: OR 0.81 (95% CI: 0.75-0.87).

### The association between influenza vaccination and COVID-19 clinical outcomes

The results of the meta-analysis that was conducted separately for each clinical outcome of interest are shown in Table 4. The results suggest a potential association between influenza vaccination and hospitalization (OR: 0.88, 95% CI: 0.81-0.96) and mechanical ventilation/invasive respiratory support (OR: 0.73, 95% CI: 0.58-0.92), but not with ICU admission (OR: 0.96, 95% CI: 0.88-1.06) and mortality (OR: 0.90, 95% CI:0.81-1.01). Both Begg’s and Egger’s tests showed no evidence of publication bias for the pooled estimates of the association between influenza vaccination and COVID-19 related clinical outcomes (all *p*-values >0.05). The summary of the publication bias tests is presented in Table 4. Figure 3 represents the forest plot for the association between influenza vaccination and COVID-19 related hospitalization and between influenza vaccination and mortality.

**Figure 3.**
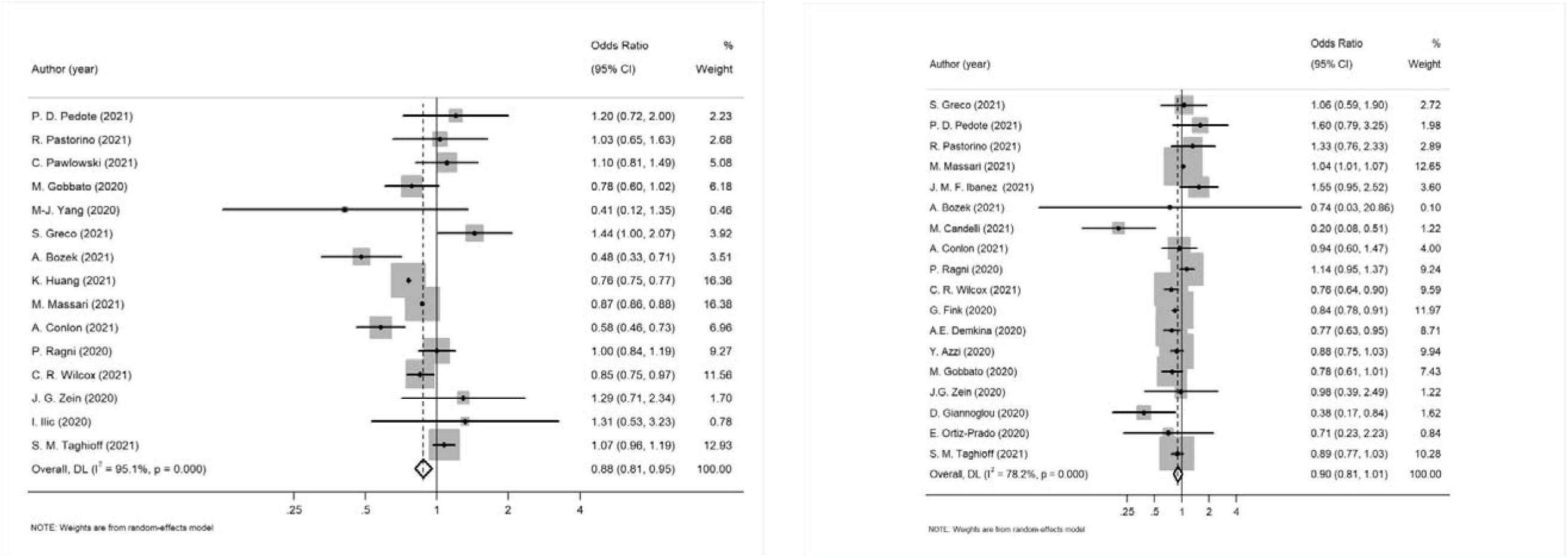
Left: Forest plot for the association between influenza vaccination and COVID-19 related hospitalization and Right: Forest plot for the association between influenza vaccination and mortality

In the sensitivity analyses that followed, the pooled estimates were consistent when any single study was omitted. Subgroup analysis was performed to explain the between studies heterogeneity for the association of influenza vaccination and COVID-19 clinical outcomes (Table 6). Study design was not used as a variable in this subgroup analysis, as the majority of the studies were cohort studies with the exception of one cross-sectional study in the hospitalization outcome and two cross-sectional study investigating the mortality outcome. Study population was also not used as the majority of the studies involved adults from the general population. Moreover, due to the limited studies reporting adjusted measures of the effect of the association between influenza vaccination and the need of mechanical ventilation, subgroup analysis could not be performed in this case.

**Table 6.** Subgroup analysis of the association between influenza vaccination and COVID-19 clinical outcomes.

| Subgroup variables | No. of studies | Adjusted estimate<br>(95% CI) | p-value | I<sup>2</sup> value<br>(%) |
| --- | --- | --- | --- | --- |
| **HOSPITALIZATION** |  |  |  |  |
| **Effect Size** |  |  |  |  |
| OR | 10 | 0.92 (0.78-1.02) | 0.40 | 86.6 |
| RR/HR | 5 | 0.87 (0.71-0.99) | 0.03 | 71.0 |
| **Adjusted Factors** |  |  |  |  |
| age/sex/age and sex | 1 | 1.44 (1.00-2.07) | 0.05 | 89.8 |
| age, sex and comorbidities | 7 | 0.85 (0.71-1.03) | 0.49 | 66.3 |
| **COVID-19 test** |  |  |  |  |
| rt-PCR | 10 | 0.89 (0.78-1.02) | 0.09 | 74.6 |
| Laboratory confirmed/(not specified) | 5 | 0.88 (0.67-1.12) | 0.30 | 90.0 |
| **Continent** |  |  |  |  |
| Europe | 10 | 0.93 (0.83-1.04) | 0.21 | 75.7 |
| N. America | 5 | 0.80 (0.63-1.02) | 0.07 | 73.1 |
| **INTENSIVE CARE UNIT** |  |  |  |  |
| **Effect Size** |  |  |  |  |
| OR | 9 | 0.91 (0.76-1.08) | 0.26 | 55.9 |
| HR | 2 | 1.01 (0.99-1.04) | 0.43 | 0.0 |
| **COVID-19 test** |  |  |  |  |
| rt-PCR | 9 | 0.93 (0.84-1.03) | 0.14 | 58.5 |
| Laboratory confirmed/(not specified) | 2 | 1.12 (0.80-1.57) | 0.52 | 14.1 |
| **Adjusted Factors** |  |  |  |  |
| age, sex and comorbidities | 4 | 0.87 (0.65-1.17) | 0.35 | 17.6 |
| **Continent** |  |  |  |  |
| Europe | 5 | 1.05 (0.95-1.16) | 0.32 | 20.6 |
| N. America | 4 | 0.69 (0.47-1.02) | 0.06 | 10.3 |
| S. America | 1 | 0.93 (0.87-0.99) | 0.02 | - |
| Asia | 1 | 0.76 (0.59-0.98) | 0.03 | - |
| **MORTALITY** |  |  |  |  |
| **Effect Size** |  |  |  |  |
| OR | 12 | 0.87 (0.76-0.99) | 0.03 | 60.5 |
| HR | 6 | 0.99 (0.86-1.12) | 0.82 | 46.3 |
| **COVID-19 test** |  |  |  |  |
| rt-PCR | 14 | 0.93 (0.82-1.06) | 0.27 | 80.3 |
| Laboratory confirmed/(not specified) | 4 | 0.81 (0.65-1.00) | 0.05 | 37.1 |
| **Adjusted Factors** |  |  |  |  |
| age/sex/age and sex | 3 | 1.07 (0.75-1.53) | 0.70 | 59.2 |
| age, sex and comorbidities | 8 | 0.82 (0.61-1.10) | 0.19 | 75.3 |
| **Continent** |  |  |  |  |
| Europe | 12 | 0.94 (0.80-1.09) | 0.40 | 75.3 |
| N. America | 3 | 0.89 (0.76-1.03) | 0.12 | 0.0 |
| S. America | 2 | 0.84 (0.78-0.91) | <0.01 | 0.0 |
| Asia | 1 | 0.78 (0.63-0.95) | 0.02 | - |

We also investigated the effect of the study of Greco et al. [38], in which they were significant differences in terms of the age of the vaccinated and non-vaccinated group and the two groups were adjusted only for sex. When the study was excluded from the meta-analysis concerning the association between influenza vaccination and mortality, the results did not significantly differ from the ones in the overall meta-analysis (OR 0.90, 95% CI: 0.81-1.00).

### The association between pneumococcal vaccination with SARS-CoV-2 infection and its clinical outcomes

A meta-analysis of the association between pneumococcal vaccination with SARS-CoV-2 infection, the need of hospitalization and ICU admission was performed. As a result, pneumococcal vaccination was shown to be associated with lower risk of COVID-19 infection (Figure 4, Table 7). The majority of the studies here (7 out of 10) provided estimates adjusted for age, sex, comorbidities and some indices of socioeconomic status (like residence, income or education). Even when we restricted the analysis in this sample the overall effect did not change (OR: 0.61, 95% CI: 0.44-0.85).

**Figure 4.**
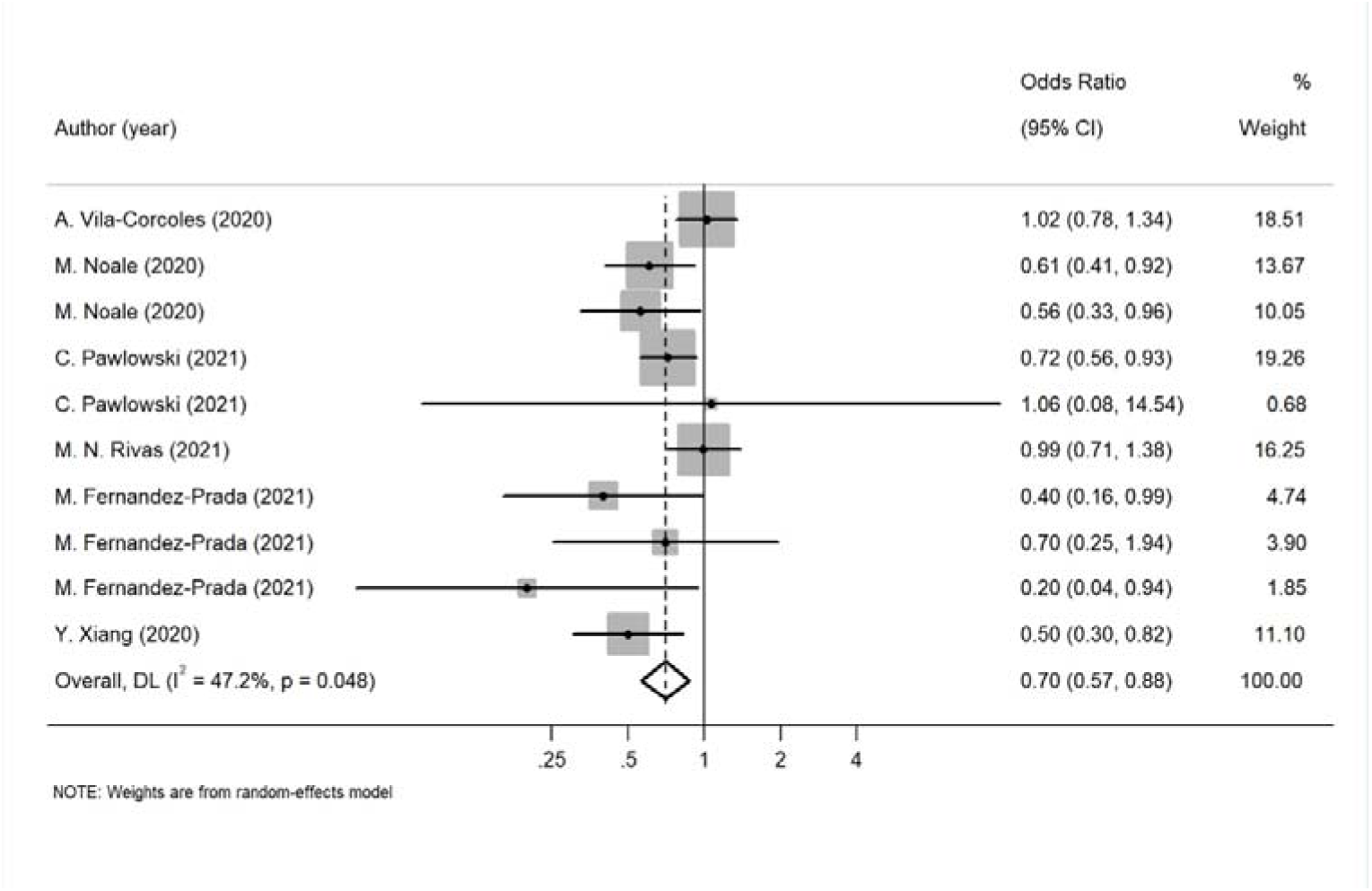
Forest plot for the association between pneumococcal vaccination and SARS-CoV-2 infection

**Table 7.** Effect sizes, heterogeneity statistics and results for publications bias of the association between pneumococcal vaccination and SARS-CoV-2 infection and outcomes.

| **Outcomes** | **No. of studies** | **Adjusted estimates (95% CI)** | ***p*-value** | ***I*<sup>2</sup> value (%)** | **Bias (*p*-value)** |
| --- | --- | --- | --- | --- | --- |
| SARS-Co V-2 infection | 10 | 0.70 (0.57-0.88) | <0.01 | 47.2 | Begg's test: 0.37<br>Egger's test: 0.11 |
| Hospitalization | 4 | 1.47 (1.30-1.67) | <0.01 | 11.4 | Begg's test: 0.09<br>Egger's test: 0.05 |
| Intensive care unit admission | 3 | 0.99 (0.60- 1.64) | 0.96 | 0.0 | Begg's test: 0.30<br>Egger's test: 0.30 |

The results of the meta-analysis regarding pneumococcal vaccination and the need of hospitalization showed that the vaccine seems to be associated with a higher risk for COVID-19 hospitalization. However, only four studies investigated the particular association, and additionally there was evidence for publication bias in this analysis making the results questionable. Finally, pneumococcal vaccination was not associated with the need of intensive care but as before, only three studies investigated this association.

## DISCUSSION

Influenza and COVID-19 are different respiratory viral diseases that can be clinically indistinguishable and co-exist at the same period. Some preliminary studies suggest some protection against SARS-CoV-2 to be conferred from vaccination to other pathogens such as influenza [14] while others have produced contrasting result [39]. There is also a limited number of studies that examined the association of the pneumococcal vaccine with the risk of developing SARS-CoV-2 infection and disease severity or risk of death on COVID-19 patients. In order to elucidate this discrepancy, we conducted a systematic review and meta-analysis to assess the association between seasonal influenza vaccination, pneumococcal vaccination and SARS-CoV-2 infection as well as with clinical outcomes related to COVID-19 morbidity and mortality. Our study followed all the available guidelines and ultimately included 38 publications that have evaluated the association of interest (24 studies on SARS-CoV-2 infection). Overall, our results indicate that influenza vaccination seems to be associated with lower risk for COVID-19 infection, as well as against hospitalization and mechanical ventilation, but no significant effect was found against “harder” endpoints like ICU admission and mortality. Specifically, the results of the meta-analysis showed that people vaccinated between autumn 2019 to autumn 2020, had up to 20% lower risk of COVID-19 infection. Similarly, if vaccinated individuals were infected by SARS-CoV-2 the risk of hospitalization would be reduced by 12% and the need of mechanical ventilation by 27%. As far as the pneumococcal vaccination is concerned, the results show that it has a protective effect against SARS-CoV-2 infection while it does not seem to have the same effect on COVID-19 hospitalization or ICU admission, although these analyses included a small number of studies.

During the last year, two systematic review and meta-analyses were published or the same topic, but the included studies were fewer that the ones included in our meta-analysis [40, 41]. In the study of Wang et al. a total of 9, 3, 2, and 3 studies were included for the association of influenza vaccination and SARS-CoV-2 infection, hospitalization, ICU admission and mortality respectively. The authors stated in their results that “the association between influenza vaccination and COVID-19 clinical outcomes was not statistically significant by random effects model while the results by fixed effects model was somehow significant”. They attribute this result to “the substantial heterogeneity between the small number of studies and participants involved in each outcome”. In their meta-analysis, Zeynali Bujani et al. [41] included 9 studies for the association of influenza vaccine and COVID-19, among which were two studies excluded from our meta-analysis. The particular studies, of Jedi et al. [42] and Caban-Marntinez et al. [43], were excluded as they reported effect sizes based on crude counts. Such data clearly violated our inclusion criteria, as we attempted to minimize the potential confounding by using only adjusted estimates. In addition, the study of Skowronski et al. [44] a retrospective analysis from Canada, which involved specimens collected during the 2010-2011 to 2016-2017 seasons, when specimens were tested for both influenza and non-influenza respiratory viruses (NIRVs), including some seasonal coronaviruses but not specific and exclusively SARS-CoV-2. We performed the meta-analysis following strict criteria, we included all the available data and investigated every study level covariate that may have influenced the outcome. Nevertheless, the between studies heterogeneity was high and the attempts to explain it using study-level covariates did not reveal anything of importance. In any case, heterogeneity is expected for many reasons that include the design of the study, the studied population, the sampling method, as well as the testing and vaccination policies in each country. In our analysis we included studies in which the seasonal influenza vaccination was administered between 2019 and 2020. The type of influenza vaccination and the exact date when the vaccines were administered was not specified in all studies and as influenza varies across years, results should be interpreted with caution regarding different influenza seasons. The testing policy implemented in each country also varied in the included studies, using different diagnostic tests. Most of the studies used molecular diagnosis with rt-PCR, few used antibody testing whereas some studies did not specify the laboratory method used. Furthermore, we must consider that national influenza immunization policies can vary significantly from country to country. These differences arise from insufficient information of the relevance of influenza infection from a clinical, social and economic point of view [45].

Our findings suggest that both vaccines are associated with a lower risk for SARS-CoV-2 infection, but not with “harder” endpoints such as ICU admission and death. The influenza vaccine was found additionally to be associated with decreased risk for hospitalization and need for mechanical ventilation. This association is counter-intuitive at first sight and hence needs to be explained. It is worth mentioning that BCG vaccine, a vaccine primarily used against tuberculosis (TB), which is mandatory in several countries [30], seems also to has a protective effect since a meta-analysis of data from 160 countries has shown that when the BCG vaccine coverage was over 70% there was a reduction in the COVID-19 infections [12]. Also some studies have found that mandatory BCG vaccination is associated with a flattening of the curve in the spread of COVID-19 [46] and that differences in mortality produced by COVID-19 across countries are correlated with a country’s BCG vaccination policy [47, 48]. However, these data come from ecological studies and thus need to be interpreted with caution.

Cross-reactivity is unlikely to exist between SARS-CoV-2 and influenza viruses, and thus other potential explanations should be considered. There is, for instance, some evidence that vaccines developed for a specific pathogen may have a wider role in protection against unrelated pathogens. Such cases include Bacillus Calmette-Guérin (BCG), measles and influenza [49]. The underlying mechanisms for such non-specific actions remain poorly understood, but a plausible suggested mechanism includes the induction of innate immune response following live attenuated vaccination, in a way that is independent of memory cells (T or B cells). This type of immunity, termed “trained innate immunity”, may confer non-specific protection against different pathogens, by inducing upregulation of recognition receptors (such as toll-like receptors) and secretion of proinflammatory cytokines (such as TNF-alpha and IL-6) in peripheral blood leucocytes and functional changes in natural killer cells [37, 50].

Nevertheless, other more plausible explanations should not be ruled out. The studies included in this meta-analysis are heterogeneous in many respects including their design, the study population and the inclusion criteria. In terms of their design all included studies were observational, so the risk of possible confounding should be considered. In such cases, when some important risk factors do exist (such as age, gender, and comorbidities), it would be unwise to perform the meta-analysis including crude estimates. However, even though we included only adjusted estimates, or studies with matched groups, the factors for which the adjustment were performed were not the same across studies. For instance, most studies adjusted for age and sex, others for several comorbidities (and even in this case, not for the same comorbidities), and some others for additional factors such as area of residence or socioeconomic status. In the subgroup analysis we tried to investigate this effect, but we were unable to find systematic differences across studies.

Socioeconomic factors, such as educational level, household size and income have been reported to have a significant effect on the probability of being vaccinated against influenza [51], whereas the same factors have been consistently shown to play significant role in COVID-19 morbidity and mortality [52–54]. It is possible therefore to speculate that individuals who get vaccinated may pay more attention to their health status and lifestyle, and subsequently, they might have been more compliant to the various non-pharmaceutical interventions suggested for COVID-19 prevention, such as social distancing, wearing masks, hand hygiene practices and use of protective equipment, leading to a reduced potential risk of infection. Following the same rationale, since vaccinated individuals are under-represented in the lower socioeconomic categories (e.g. those who cannot work from home, those who need to use public transportation, or live in crowded households and so on), then these factors might explain the observed increased risk of getting COVID-19. This interpretation is also consistent with the results suggesting that vaccinated individuals are at increased risk of SARS-CoV-2 infection, but not at risk of ICU admission or death, since in these cases the comparison was made among those that were already infected, and the supposed confounding effect has been potentially removed.

Interestingly, when the analysis was restricted to the studies that adjusted for age, sex, comorbidities and some indices of socioeconomic status (like area of residence, income or education), the association of influenza vaccination with the risk of SARS-CoV-2 infection was diminished. Thus, future and more carefully designed studies, that will adjust more thoroughly for socioeconomic indices, should be pursuit to investigate this hypothesis. However, this observation does not seem to hold regarding the association of pneumococcal vaccination with SARS-CoV-2 infection, since the statistically significant association persists even after restricting the analysis in the studies that adjusted for the same factors. We need to mention, that even though the studies regarding pneumococcal vaccination were fewer in total, the majority of them provided estimates adjusted for socioeconomic indices, which is in contrast to what is the case for studies of influenza vaccine.

Most respiratory viral diseases may have as complication bacterial coinfections, or secondary infections (super-infections), which can worsen the clinical outcome increasing thus morbidity and mortality. However, it has been argued that the proportion of COVID-19 patients with bacterial coinfections and/or super-infections may be lower compared to what we have seen in patients suffering from influenza [55–57]. Nevertheless, since vaccination is beneficial per se, it has been suggested that pneumococcal vaccination can, to some extent, provide additional protection to the COVID-19 patients reducing the morbidity and mortality [58]. Animal studies have also shown that an initial SARS-CoV-2 infection can increase susceptibility and pathogenicity to bacterial coinfection [59]. The limited number of available studies regarding the association of pneumococcal vaccination with hospitalization, ICU admission and death, did not allow us to test this hypothesis, but as we said the association of vaccination with SARS-CoV-2 infection remained significant even after adjustment for sex, age, comorbidities and socioeconomic indices, and this should be investigated in future studies.

## Conflict of Interest

none declared

## Data Availability

All data produced in the present study are available upon reasonable request to the authors

## Notes

### Competing Interest Statement

The authors have declared no competing interest.

### Funding Statement

This study did not receive any funding

## REFERENCES

1. Huang, C., et al., Clinical features of patients infected with 2019 novel coronavirus in Wuhan, China. The lancet, 2020. 395(10223): p. 497–506.

2. Maier, H.J., E. Bickerton, and P. Britton, Preface. Coronaviruses. Methods Mol Biol, 2015. 1282: p. v.

3. Lu, R., et al., Genomic characterisation and epidemiology of 2019 novel coronavirus: implications for virus origins and receptor binding. Lancet (London, England), 2020. 395(10224): p. 565–574.

4. (WHO), W.H.O. Origin of SARS-CoV-2. 2019; WHO/2019-nCoV/FAQ/Virus_origin/2020.1].

5. Bouvier, N.M. and P. Palese, The biology of influenza viruses. Vaccine, 2008. 26 Suppl 4(Suppl 4): p. D49–53.

6. (WHO), W.H.O. Influenza (Seasonal). 2018.

7. Jiang, C., et al., Comparative review of respiratory diseases caused by coronaviruses and influenza A viruses during epidemic season. Microbes Infect, 2020. 22(6-7): p. 236–244.

8. (WHO), W.H.O. Pneumonia. 2021.

9. Xing, Q.-s., et al., Precautions are Needed for COVID-19 Patients with Coinfection of Common Respiratory Pathogens. medRxiv, 2020: p. 2020.02.29.20027698.

10. Team, V.G.C.V.T. COVID-19 Vaccine Tracker 2022; Available from: https://covid19.trackvaccines.org/.

11. Amirlak, L., et al., Effectiveness of booster BCG vaccination in preventing Covid-19 infection. Hum Vaccin Immunother, 2021: p. 1–3.

12. Joy, M., et al., Is BCG associated with reduced incidence of COVID-19? A meta-regression of global data from 160 countries. Clinical epidemiology and global health, 2021. 9: p. 202–203.

13. Dolgikh, S., Hospitalization Data Supports Correlation of Lower Covid-19 Severity vs. Universal BCG Immunization in the Early Phase of the Pandemic. medRxiv, 2021.

14. Mosaddeghi, P., et al., Harnessing the non-specific immunogenic effects of available vaccines to combat COVID-19. Hum Vaccin Immunother, 2021. 17(6): p. 1650–1661.

15. Kissling, E., et al., Absence of association between 2019-20 influenza vaccination and COVID-19: Results of the European I-MOVE-COVID-19 primary care project, March-August 2020. Influenza Other Respir Viruses, 2021. 15(4): p. 429–438.

16. PubMed Help [Internet], in Bethesda (MD): National Center for Biotechnology Information (US); 2005-. PubMed Help. [Updated 2018 Mar 28].

17. medRxiv. 2019.

18. Moher, D., et al., Preferred reporting items for systematic reviews and meta-analyses: the PRISMA statement. PLoS Med, 2009. 6(7): p. e1000097.

19. Morton, S.C., et al., Standards and guidelines for observational studies: quality is in the eye of the beholder. Journal of Clinical Epidemiology, 2016. 71: p. 3–10.

20. Forero, D.A., et al., Ten simple rules for carrying out and writing meta-analyses. PLoS computational biology, 2019. 15(5): p. e1006922–e1006922.

21. DerSimonian, R. and N. Laird, Meta-analysis in clinical trials. Control Clin Trials, 1986. 7(3): p. 177–88.

22. Higgins, J.P., et al., Measuring inconsistency in meta-analyses. Bmj, 2003. 327(7414): p. 557–60.

23. Begg, C.B. and M. Mazumdar, Operating characteristics of a rank correlation test for publication bias. Biometrics, 1994. 50(4): p. 1088–101.

24. Egger, M., et al., Bias in meta-analysis detected by a simple, graphical test. Bmj, 1997. 315(7109): p. 629–34.

25. StataCorp, Stata 13 Base Reference Manual. 2013, College Station, TX: Stata Press.

26. Martinez-Baz, I., et al., Influenza Vaccination and Risk of SARS-CoV-2 Infection in a Cohort of Health Workers. Vaccines (Basel), 2020. 8(4).

27. Massoudi, N. and B. Mohit, A Case-Control Study of the 2019 Influenza Vaccine and Incidence of COVID-19 Among Healthcare Workers. J Clin Immunol, 2021. 41(2): p. 324–334.

28. Belingheri, M., et al., Association between seasonal flu vaccination and COVID-19 among healthcare workers. Occup Med (Lond), 2020. 70(9): p. 665–671.

29. Rivas, M.N., et al., BCG vaccination history associates with decreased SARS-CoV-2 seroprevalence across a diverse cohort of health care workers. J Clin Invest, 2021. 131(2).

30. Erismis, B., et al., Annual influenza vaccination effect on the susceptibility to COVID-19 infection. Cent Eur J Public Health, 2021. 29(1): p. 14–17.

31. de la Cruz Conty, M.L., et al., Impact of Recommended Maternal Vaccination Programs on the Clinical Presentation of SARS-CoV-2 Infection: A Prospective Observational Study. Vaccines (Basel), 2021. 9(1).

32. Azzi, Y., et al., COVID-19 infection in kidney transplant recipients at the epicenter of pandemics. Kidney Int, 2020. 98(6): p. 1559–1567.

33. Caratozzolo, S., et al., The impact of COVID-19 on health status of home-dwelling elderly patients with dementia in East Lombardy, Italy: results from COVIDEM network. Aging Clin Exp Res, 2020. 32(10): p. 2133–2140.

34. Huang, K., et al., Influenza vaccination and the risk of COVID-19 infection and severe illness in older adults in the United States. Sci Rep, 2021. 11(1): p. 11025.

35. Noale, M., et al., The Association between Influenza and Pneumococcal Vaccinations and SARS-Cov-2 Infection: Data from the EPICOVID19 Web-Based Survey. Vaccines (Basel), 2020. 8(3).

36. Pastorino, R., et al., Influenza and pneumococcal vaccinations are not associated to COVID-19 outcomes among patients admitted to a university hospital. Vaccine, 2021. 39(26): p. 3493–3497.

37. Debisarun, P.A., et al., The effect of influenza vaccination on trained immunity: impact on COVID-19. MedRxiv, 2020.

38. Greco, S., et al., SARS-CoV-2 infection and H1N1 vaccination: does a relationship between the two factors really exist? A retrospective analysis of a territorial cohort in Ferrara, Italy. Eur Rev Med Pharmacol Sci, 2021. 25(6): p. 2795–2801.

39. Kissling, E., et al., Absence of association between 2019-20 influenza vaccination and COVID-19: Results of the European I-MOVE-COVID-19 primary care project, March-August 2020. Influenza and other respiratory viruses, 2021. 15(4): p. 429–438.

40. Wang, R., M. Liu, and J. Liu, The Association between Influenza Vaccination and COVID-19 and Its Outcomes: A Systematic Review and Meta-Analysis of Observational Studies. Vaccines (Basel), 2021. 9(5).

41. Zeynali Bujani, M., et al., The Effect of Influenza Vaccination on COVID-19 Morbidity, Severity and Mortality: Systematic Review and Meta-Analysis. Malays J Med Sci, 2021. 28(6): p. 20–31.

42. Jehi, L., et al., Individualizing Risk Prediction for Positive Coronavirus Disease 2019 Testing: Results From 11,672 Patients. Chest, 2020. 158(4): p. 1364–1375.

43. Caban-Martinez, A.J., et al., Epidemiology of SARS-CoV-2 antibodies among firefighters/paramedics of a US fire department: a cross-sectional study. Occup Environ Med, 2020. 77(12): p. 857–861.

44. Skowronski, D.M., et al., Influenza Vaccine Does Not Increase the Risk of Coronavirus or Other Noninfluenza Respiratory Viruses: Retrospective Analysis From Canada, 2010-2011 to 2016-2017. Clin Infect Dis, 2020. 71(16): p. 2285–2288.

45. Principi, N., et al., Influenza immunization policies: Which could be the main reasons for differences among countries? Human vaccines & immunotherapeutics, 2018. 14(3): p. 684–692.

46. Berg, M.K., et al., Mandated Bacillus Calmette-Guérin (BCG) vaccination predicts flattened curves for the spread of COVID-19. Science advances, 2020. 6(32): p. eabc1463.

47. Miller, A., et al., Correlation between universal BCG vaccination policy and reduced mortality for COVID-19. MedRxiv, 2020.

48. Singh, S., et al., BCG vaccination impact on mortality and recovery rates in COVID-19: A meta-analysis. Monaldi Archives for Chest Disease, 2021.

49. Covián, C., et al., BCG-induced cross-protection and development of trained immunity: implication for vaccine design. Frontiers in Immunology, 2019. 10: p. 2806.

50. Long, B.R., et al., Elevated frequency of gamma interferon-producing NK cells in healthy adults vaccinated against influenza virus. Clinical and Vaccine Immunology, 2008. 15(1): p. 120–130.

51. Endrich, M.M., P.R. Blank, and T.D. Szucs, Influenza vaccination uptake and socioeconomic determinants in 11 European countries. Vaccine, 2009. 27(30): p. 4018–4024.

52. Rollston, R. and S. Galea, COVID-19 and the social determinants of health. 2020, SAGE Publications Sage CA: Los Angeles, CA. p. 687–689.

53. Patel, J., et al., Poverty, inequality and COVID-19: the forgotten vulnerable. Public health, 2020. 183: p. 110.

54. Wachtler, B., et al., Socioeconomic inequalities in the risk of SARS-CoV-2 infection– First results from an analysis of surveillance data from Germany. 2020.

55. Lansbury, L., et al., Co-infections in people with COVID-19: a systematic review and meta-analysis. Journal of Infection, 2020. 81(2): p. 266–275.

56. Feldman, C. and R. Anderson, The role of co-infections and secondary infections in patients with COVID-19. Pneumonia, 2021. 13(1): p. 1–15.

57. Joseph, C., Y. Togawa, and N. Shindo, Bacterial and viral infections associated with influenza. Influenza and other respiratory viruses, 2013. 7: p. 105–113.

58. Im, H., et al., Promising Expectations for Pneumococcal Vaccination during COVID-19. Vaccines, 2021. 9(12): p. 1507.

59. Smith, A.P., et al., Time-Dependent Increase in Susceptibility and Severity of Secondary Bacterial Infection during SARS-CoV-2 Infection. bioRxiv, 2022.

60. Ragni, P., et al., Association between Exposure to Influenza Vaccination and COVID-19 Diagnosis and Outcomes. Vaccines (Basel), 2020. 8(4).

61. Vila-Corcoles, A., et al., Influence of prior comorbidities and chronic medications use on the risk of COVID-19 in adults: a population-based cohort study in Tarragona, Spain. BMJ Open, 2020. 10(12): p. e041577.

62. Green, I., et al., The association of previous influenza vaccination and coronavirus disease-2019. Hum Vaccin Immunother, 2021. 17(7): p. 2169–2175.

63. Conlon, A., et al., Impact of the influenza vaccine on COVID-19 infection rates and severity. Am J Infect Control, 2021. 49(6): p. 694–700.

64. Bozek, A., et al., Impact of influenza vaccination on the risk of SARS-CoV-2 infection in a middle-aged group of people. Hum Vaccin Immunother, 2021. 17(9): p. 3126–3130.

65. Kowalska, M., et al., Association between Influenza Vaccination and Positive SARS-CoV-2 IgG and IgM Tests in the General Population of Katowice Region, Poland. Vaccines (Basel), 2021. 9(5).

66. Fernandez-Prada, M., et al., Personal and vaccination history as factors associated with SARS-CoV-2 infection. Med Clin (Engl Ed), 2021. 157(5): p. 226–233.

67. King, J.P., H.Q. McLean, and E.A. Belongia, Risk of symptomatic severe acute respiratory syndrome coronavirus 2 infection not associated with influenza vaccination in the 2019-2020 season. Influenza Other Respir Viruses, 2021. 15(6): p. 697–700.

68. Pawlowski, C., et al., Exploratory analysis of immunization records highlights decreased SARS-CoV-2 rates in individuals with recent non-COVID-19 vaccinations. Sci Rep, 2021. 11(1): p. 4741.

69. Zein, J.G., G. Whelan, and S.C. Erzurum, Safety of influenza vaccine during COVID-19. Journal of Clinical and Translational Science, 2021. 5(1).

70. Xiang, Y., K.C. Wong, and H.C. So, Exploring Drugs and Vaccines Associated with Altered Risks and Severity of COVID-19: A UK Biobank Cohort Study of All ATC Level-4 Drug Categories Reveals Repositioning Opportunities. Pharmaceutics, 2021. 13(9).

71. Alkathlan, M., et al., Trends, Uptake, and Predictors of Influenza Vaccination Among Healthcare Practitioners During the COVID-19 Pandemic Flu Season (2020) and the Following Season (2021) in Saudi Arabia. J Multidiscip Healthc, 2021. 14: p. 2527–2536.

72. Pedote, P.D., et al., Influenza Vaccination and Health Outcomes in COVID-19 Patients: A Retrospective Cohort Study. Vaccines (Basel), 2021. 9(4).

73. Gobbato, M., et al., Clinical, demographical characteristics and hospitalisation of 3,010 patients with Covid-19 in Friuli Venezia Giulia Region (Northern Italy). A multivariate, population-based, statistical analysis. Epidemiol Prev, 2020. 44(5-6 Suppl 2): p. 226–234.

74. Yang, M.J., et al., Influenza Vaccination and Hospitalizations Among COVID-19 Infected Adults. J Am Board Fam Med, 2021. 34(Suppl): p. S179–S182.

75. Massari, M., et al., Association of Influenza Vaccination and Prognosis in Patients Testing Positive to SARS-CoV-2 Swab Test: A Large-Scale Italian Multi-Database Cohort Study. Vaccines (Basel), 2021. 9(7).

76. Wilcox, C.R., N. Islam, and H. Dambha-Miller, Association between influenza vaccination and hospitalisation or all-cause mortality in people with COVID-19: a retrospective cohort study. BMJ Open Respir Res, 2021. 8(1).

77. Ilic, I., et al., Pneumonia in medical professionals during COVID-19 outbreak in cardiovascular hospital. Int J Infect Dis, 2021. 103: p. 188–193.

78. Taghioff, S.M., et al., Examining the potential benefits of the influenza vaccine against SARS-CoV-2: A retrospective cohort analysis of 74,754 patients. PLoS One, 2021. 16(8): p. e0255541.

79. Fink, G., et al., Inactivated trivalent influenza vaccination is associated with lower mortality among patients with COVID-19 in Brazil. BMJ Evid Based Med, 2020.

80. Demkina, A.E., et al., Risk factors for outcomes of COVID-19 patients: an observational study of 795 572 patients in Russia. medRxiv, 2020.

81. Candelli, M., et al., Effect of influenza vaccine on COVID-19 mortality: A retrospective study. Internal and emergency medicine, 2021. 16(7): p. 1849–1855.

82. Fernandez Ibanez, J.M., et al., Influence of influenza vaccine and comorbidity on the evolution of hospitalized COVID-19 patients. Med Clin (Barc), 2021.

83. Giannoglou, D., et al., Predictors of mortality in hospitalized COVID-19 patients in Athens, Greece. medRxiv, 2020.

84. Ortiz-Prado, E., et al., Epidemiological, socio-demographic and clinical features of the early phase of the COVID-19 epidemic in Ecuador. PLoS Negl Trop Dis, 2021. 15(1): p. e0008958.

